# Operationalizing a routine wastewater monitoring laboratory for SARS-CoV-2

**DOI:** 10.1101/2021.06.06.21258431

**Authors:** Rose S. Kantor, Hannah D. Greenwald, Lauren C. Kennedy, Adrian Hinkle, Sasha Harris-Lovett, Matthew Metzger, Melissa M. Thornton, Justin M. Paluba, Kara L. Nelson

## Abstract

Wastewater-based testing for SARS-CoV-2 is a novel tool for public health monitoring, but additional laboratory capacity is needed to provide routine monitoring at all locations where it has the potential to be useful. Few standardization practices for SARS-CoV-2 wastewater analysis currently exist, and quality assurance/quality control procedures may vary across laboratories. Alongside counterparts at many academic institutions, we built out a laboratory for routine monitoring of wastewater at the University of California, Berkeley. Here, we detail our group’s establishment of a wastewater testing laboratory including standard operating procedures, laboratory buildout and workflow, and a quality assurance plan. We present a complete data analysis pipeline and quality scoring framework and discuss the data reporting process. We hope that this information will aid others at research institutions, public health departments, and wastewater agencies in developing programs to support wastewater monitoring for public health decision-making.

## 1. Introduction

Wastewater-based epidemiology (WBE) is a pandemic response tool that provides population-level public health information to complement clinical testing and other epidemiological data^1–5^. At the outset of the coronavirus disease 2019 (COVID-19) pandemic, academic, commercial, and wastewater utility laboratories developed and optimized protocols for concentration, extraction, and quantification of SARS-CoV-2 RNA from wastewater^1,2,5–14^. Due to an unprecedented level of collaboration,^15^ standard operating procedures (SOPs) and commercial kits now enable routine wastewater monitoring. However, in many locales, implementation of WBE is still in the early stages or is housed in academic research laboratories^16^.

As wastewater monitoring enters a new phase with the implementation of the Center for Disease Control and Prevention’s National Wastewater Surveillance System (CDC-NWSS)^17^, there may be a need to expand the capacity of existing laboratories or establish new ones. There is also an ongoing need for transparency and documentation of raw data analysis methods and quality controls. In particular, quality control for WBE analysis has not been standardized, and conveying the quality of results with multiple analytical controls is complex^18^. Finally, submitting data to CDC-NWSS requires conformation to data field code specifications which can be time-consuming to perform manually.

In October 2020, we launched a laboratory at the University of California, Berkeley, to support wastewater monitoring for SARS-CoV-2 in the San Francisco Bay Area. We developed and optimized standard operating procedures, workflows, and a data analysis code base. The main contributions of this work are to provide: 1) a start-to-finish guidance document on laboratory set-up & operation drawn from our experience, 2) a data analysis pipeline for WBE that implements best practices in the field and is compatible with CDC-NWSS, 3) a unique data quality scoring method that can be adapted to meet the needs of other labs using different methodologies. While we recognize that there is no single preferred method or set of methods for analyzing SARS-CoV-2 in wastewater, we hope that these resources will be useful to other groups regardless of which methods are used.

## 2. Methods

Our experience is based on analyzing wastewater samples collected from residential buildings, sub-sewersheds, and influent to wastewater treatment plants using the 4S direct extraction method^19^ and quantification by qRT-PCR^20^. Nonetheless, in each section below we describe the approach in a manner that is transferable to other laboratory methods.

### 2.1 Laboratory safety and space

Current CDC guidelines for protocols involving SARS-CoV-2 genetic material (RNA) and for concentrating SARS-CoV-2 from raw wastewater or primary sludge specify that work must be conducted at biosafety level 2, with biosafety level 3 precautions for methods that concentrate virus presumed to be intact^21^. Raw wastewater samples should be shipped as biohazard class B (UN 3373)^22,23^. To minimize risk from handling raw wastewater, the 4S method for direct RNA extraction from wastewater includes minimal sample handling prior to heat-inactivation and does not involve concentration of viral particles^19^. An example Biological Use Authorization for the 4S method followed by RT-qPCR is included in the Supplementary Information. Institutional guidelines dictate the laboratory’s disposal protocols for ethanol waste (generated during RNA extraction) and biohazardous waste^24^.

The laboratory space was certified as biosafety level 2, with negative pressure. An area for donning and doffing PPE was designated outside of the main laboratory workspace. To limit amplicon cross-contamination, the laboratory was designed with a one-way path from RNA extraction to PCR preparation to RT-qPCR analysis. In the main laboratory, biosafety cabinets were dedicated exclusively to RNA extraction or RT-qPCR work, and never used for both. Separate rooms were designated for preparation (of sampling kits, buffers, and sample processing materials) and for sample receiving to prevent cross-contamination and so that laboratory volunteers were not exposed to biohazardous materials. Lastly, the laboratory space needed to be large enough to accommodate physical distancing to prevent potential spread of COVID-19 between laboratory personnel. Major laboratory equipment, accessory equipment, and non-consumable supplies are listed in **Table S1** (see **Supplementary Tables**).

### 2.2 Laboratory workflow

Our overall process workflow from sampling to data interpretation is outlined in **Figure 1**, and approximate time frames and safety requirements for each step of the process are shown in **Table S2**. Reagents and costs per sample are presented in **Table S3**.

**Figure 1.**
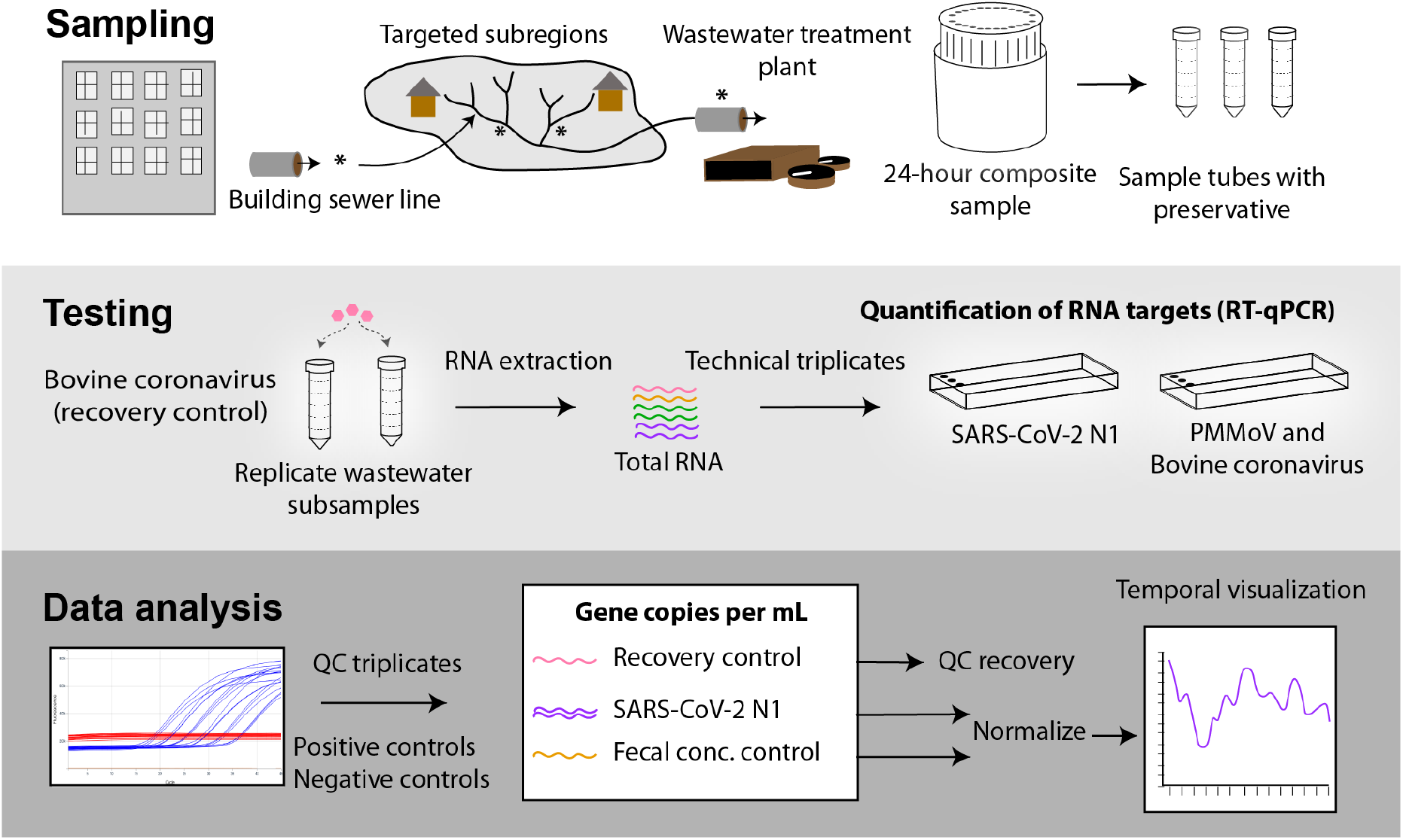
The workflow for sampling, testing, and data analysis. Twenty-four hour composite samples are collected from a building sewer line, targeted subregion, or influent to a wastewater treatment plant (denoted by asterisks). Replicate subsamples of the composite samples are spiked with bovine coronavirus and RNA is extracted (4S method). RT-qPCR assays are performed in triplicate to quantify SARS-CoV-2 (CDC-N1 target), pepper mild mottle virus (PMMoV), and bovine coronavirus (BCoV). Data analysis steps include combining RT-qPCR technical triplicates, checking standards and negative controls, and calculating gene copies per milliliter of raw wastewater.

#### Sample collection

Each week, laboratory staff prepare and ship sampling kits to wastewater agencies. The sampling kits include tubes with pre-weighed NaCl, Tris, and EDTA, such that adding the 40 mL wastewater sample results in lysis and preservation of the SARS-CoV-2 RNA^19^. The kits include instructions (**Supplementary Information**) to ensure samples are collected consistently from each agency and that associated metadata are tied to the samples. Wastewater agency personnel collect samples of raw wastewater at designated locations using 24-hour composite samplers. In general, only the entity that owns the sewer system (usually the wastewater agency or city) may enter the sewers, which means sampling is performed by or in direct collaboration with that entity. The composite sample is mixed and aliquoted into replicate sample tubes, and the tubes are returned via overnight shipping on ice to the laboratory.

#### Laboratory testing

In the lab, samples are spiked with bovine coronavirus (BCoV; Bovilis coronavirus calf vaccine, Merck) as a process control, heat inactivated, and RNA is extracted via the 4S method in batches of 19 alongside a phosphate-buffered saline negative control^19^. One sample tube from each location is temporarily stored at 4°C in case the extraction needs to be repeated. Quantification of RNA targets by RT-qPCR is performed in technical triplicate alongside a seven-point standard curve and PCR water no-template control wells^20^. The limit of detection was determined based on 18 dilution series of standard and set to the value above which 95% of the replicates amplified. To decrease inhibition, the CDC N1 assay for SARS-CoV-2 is performed on undiluted and 5-fold diluted RNA template, while the multiplexed pepper mild mottle virus (PMMoV) and BCoV assay is performed on 5-fold diluted template only. Standard operating procedures for extraction and quantification are available on protocols.io^19,20^. Regular cleaning and laboratory maintenance are performed to maintain QA/QC standards (**Supplementary Information**).

#### Data analysis

Raw data processing and QA/QC are performed using a custom pipeline (**Figure S1**; https://github.com/wastewaterlab/data_analysis). Briefly, outliers are removed from raw Cq values of qPCR triplicate wells using the Grubbs test (alpha=0.05). Next, Cq values are converted to gene copies per well based on a standard curve, substituting one-half the limit of detection for non-detects. Gene copies in triplicate wells are averaged using the geometric mean. For the N1 assay, the dilution with the higher effective quantification is reported. The concentration of the qPCR target in the raw wastewater is calculated based on the concentration factor, accounting for the initial mass of the sample and effective volume analyzed in each qPCR assay. Results are provided as SARS-CoV-2 N1 concentrations and N1 concentrations normalized to PMMoV and are visualized as temporal data series. The geometric mean and standard deviation are reported for biological replicates.

### 2.3 Laboratory inventory management system (LIMS)

In our start-up phase, validated google sheets were used as a temporary data storage system. We investigated a range of data management options including: 1) a custom relational database hosted locally, 2) a custom relational database hosted on an academic computing server, or 3) a commercial LIMS. Because commercial LIMS include a user-friendly front-end that has already been partially customized for laboratory uses, provides more security and back-ups, and ensures data validation, we chose this option. The most important elements for choosing the appropriate LIMS included: ability to communicate with the qPCR/ddPCR machine, sample barcoding, ability to include custom code for QC analysis, and an application programming interface (API) for pulling the data into a dashboard for visualization of results. Database and data analysis schematics are presented in the **Supplementary Information**.

### 2.4 Quality assurance and quality control

We use the protocols, experimental controls^25^, and replication outlined in our Quality Assurance Plan (**Supplementary Information 5**) to ensure the reliability of results. The Quality Assurance Plan represents current practices, which were implemented in phases as the laboratory’s capacity expanded. Briefly, to account for wastewater heterogeneity, wastewater samples are processed in biological replicates (i.e., 2 aliquots from the same 24-hour composite sample) with a batch extraction blank (phosphate buffered saline). To control for technical variation, each qPCR assay is run in triplicate wells for all samples, a 7-point standard curve, and controls. Lastly, the SARS-CoV-2 N1 assay is run on undiluted and 5-fold diluted template to test for and reduce inhibition. A set of minimum requirements governs decisions to rerun or exclude individual samples, extraction batches, or qPCR plates before reporting (**Supplementary Information and Table S4**).

We developed a custom scoring framework (**Table S4)** and code (https://github.com/wastewaterlab/data_analysis) to assess data quality. The framework considers all positive and negative controls as well as technical replicates and several quantitative controls. Data that fail to meet quality thresholds for efficiency, recovery, or standard conditions are flagged. Raw data and quality-scored results are reviewed by two individuals, and any issues are addressed before reporting processed data to public health agencies.

## 3. Results

### 3.1 Capacity scaling over time

Laboratory work and staff training began in a research lab, while the regional monitoring laboratory was prepared. The cost for laboratory buildout was approximately $100,000 USD in capital equipment and took 3 months to complete (**Table S1**). Upon opening in October 2020, the laboratory began processing 30 samples per week in single replicates. Sites were added at the request of local public health officials and wastewater agencies. As of May 2021, the laboratory processed 65 samples per week in biological duplicate (two subsamples per 24-hour composite sample), for a total of 130 samples per week (**Figure S2**). These samples represented 18 different wastewater agencies and 38 sites in the San Francisco Bay Area. Sites were sampled between 1 and 5 times per week, depending on wastewater agency staffing capacity. Our laboratory uses the kit-free sewage, salt, silica, and SARS-CoV-2 (4S) direct RNA extraction method^7^ and reverse transcription quantitative polymerase chain reaction (RT-qPCR) with a per-sample cost (consumables and reagents) of around $25 USD (**Table S3**). The laboratory workflow can be seen in **Figure 1**. Processing time and cost breakdowns per sample can be found in **Tables S2 and S3**.

Turnaround time, defined as the number of days between sample collection and RT-qPCR results for SARS-CoV-2 N1, averaged 1.6 days (including shipping time) in May 2021. Turnaround time from sample delivery to RT-qPCR results was typically within 24 hours. In total, at least 75% of samples had turnaround times ≤ 2 days. Holidays increased turnaround time due to slower shipping and decreased the number of samples received by the laboratory due to lower wastewater agency staffing. Initially, SARS-CoV-2 N1 was assayed for every sample, while PMMoV was assayed for one of two biological replicates and bovine coronavirus was assayed for one sample from each extraction batch. As the laboratory scaled up, PMMoV and bovine coronavirus assays were run more frequently and ultimately multiplexed, allowing them to be run on every sample.

### 3.2 Data analysis and reporting

The data analysis pipeline presented here applies common qPCR data analysis techniques such as outlier removal from technical replicates^26^, linear regression of a standard curve to convert Cq values to gene copies (**Figure S1 and Supplementary Methods**), and substitution of all values below one-half the limit of detection, including non-detects, with one-half the limit of detection (see Methods). Concentrations of SARS-CoV-2 in raw wastewater are reported as gene copies per milliliter and as normalized gene copies of SARS-CoV-2 per PMMoV. The data analysis pipeline additionally produces information that can be used to fill data fields required by CDC-NWSS (e.g. the geometric standard deviation of all qPCR replicates, for calculation of geometric standard error).

Data are reported on an internal dashboard for public health officials, where results for biological replicates are presented separately as well as combined by geometric mean with geometric standard deviation (**Figure 2**). Initially, limited sampling and processing capacity led to useful but low-resolution data. Once biological replicates were processed and/or sampling frequency increased, more of the methodological variability was captured in the data (**Figure 2**). To aid in interpretation of potential under- or over-estimation (and false negatives or false positives), data quality is also reported for each point on the dashboard (see section 3.3).

**Figure 2.**
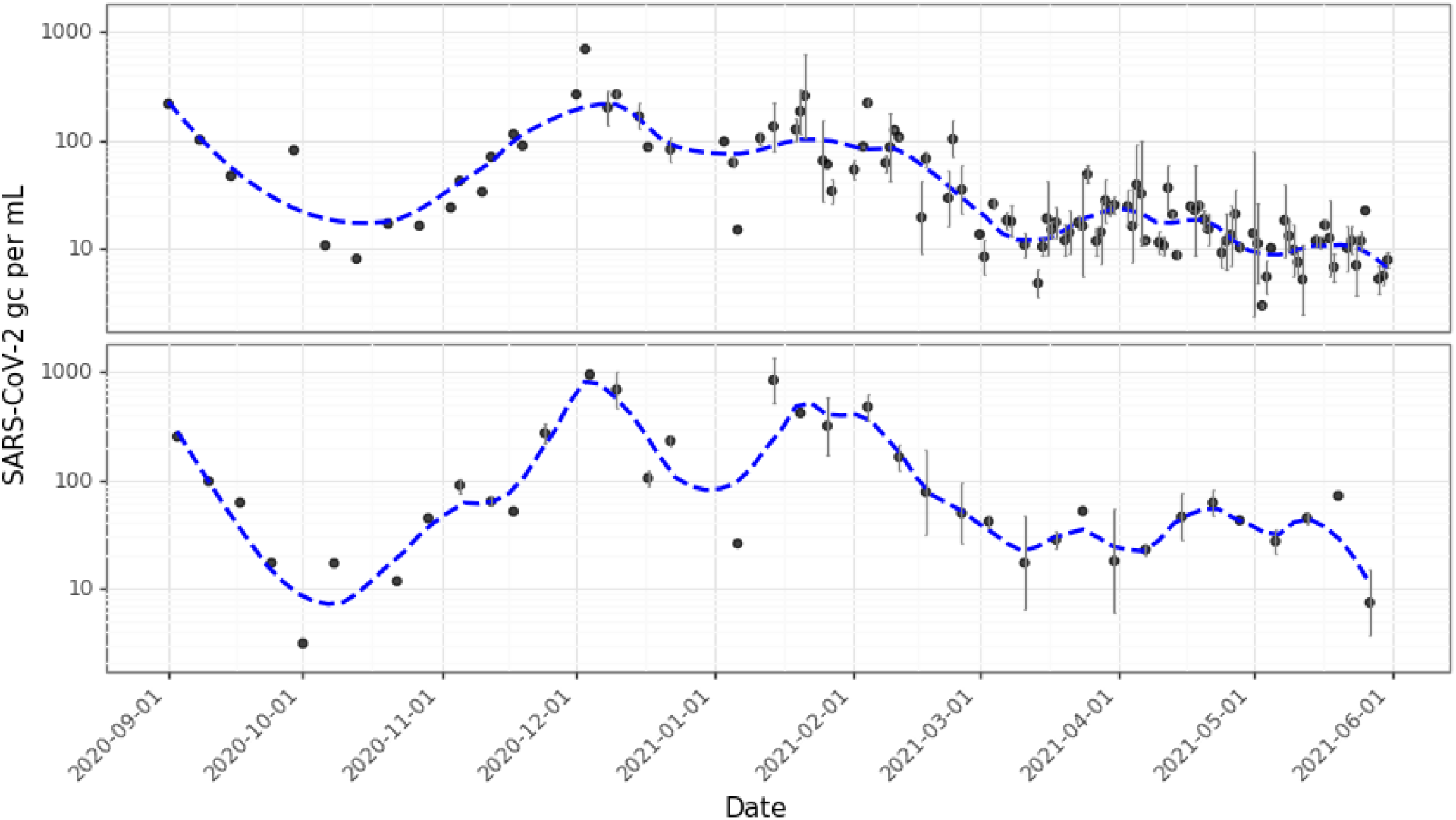
Example data for a sewershed (top) and a nested sub-sewershed (bottom) from September 2020 through mid-May 2021. The y-axis is log10-scaled SARS-CoV-2 N1 gene copies per milliliter. Where points have error bars, they are the geometric mean with geometric standard deviation of two biological replicates. The dashed blue line shows the lowess-smoothed trend (bandwidth = 0.25). For the sewershed, sample frequency increased over time: from September to December 2020, sample frequency was 1-2 times per week; from January 2020 to mid-march 2021, sample frequency was 3 times per week; after mid-March 2021, sample frequency was 5 times per week. The sub-sewershed was sampled 1-2 times per week throughout the time series. For a quality-scored version of this data showing individual biological replicates, see Figure S3.

### 3.3 Data quality and sources of variability

To convey information about the quality of each data point to data end-users, a weighted scoring framework was developed. Scoring is based on quality controls and metrics from three phases of testing: sample collection, RNA extraction, and target quantification (**Table S4**). The weighted score summarizes 11 parameters for each sample, including positive and negative controls, qualitative observations (e.g. “sample processing error”), and quantitative controls (e.g. recovery efficiency) (see **Figure S3** for scored data example). For some quantitative controls, the thresholds for poor, acceptable, and high quality were defined by theoretical ideals (e.g. qPCR efficiency should be close to 100%). In other cases, thresholds were chosen based on a meta-analysis of data from all samples (**Figures S4-S7**). Thresholds could not be easily defined statistically due to the non-normal nature of the data and were therefore assigned via visual inspection of aggregated data and laboratory experience.

Two quality parameters address potential sample cross-contamination: the extraction negative control and the no-template RT-qPCR control (see Methods; **Table S4**). The score for these controls focuses only on the SARS-CoV-2 N1 assay. In the monitoring lab, amplification of the extraction negative control in this assay was rare (4 of 249 extraction controls tested). Amplification of the extraction control in the PMMoV assay was more common (amplification in 62 of 239 extraction controls tested) at a median Cq of 37.7. We do not consider this amplification to be cause for concern because wastewater PMMoV concentrations in the San Francisco Bay Area are typically several orders of magnitude higher than this value (median Cq = 28.4, n = 2175). Several of the remaining quality parameters account for methodological factors that could contribute to variability between biological replicates or samples from consecutive days, as observed in **Figures 2** and **S3**. These include composite sampler errors, sample hold times, RNA extraction efficiency, RT-qPCR efficiency, and RT-qPCR inhibition (**Table S4**).

In a meta-analysis of factors affecting quality across all samples processed in the monitoring laboratory, the most common factors affecting the data quality were RT-qPCR efficiency and the presence of qPCR inhibition (**Figure 3**). Initially, the efficiency of the SARS-CoV-2 N1 RT-qPCR assay was lower than expected. Increased training for technicians, moving to a climate-controlled laboratory space, a shift from RNA to linear plasmid DNA standards, and pre-aliquoting weekly standard curves generally improved qPCR efficiency. Meanwhile, the introduction of new technicians resulted in shifts in efficiency and in the y-intercept (**Figures S8 and S9**; see supplementary methods).

**Figure 3.**
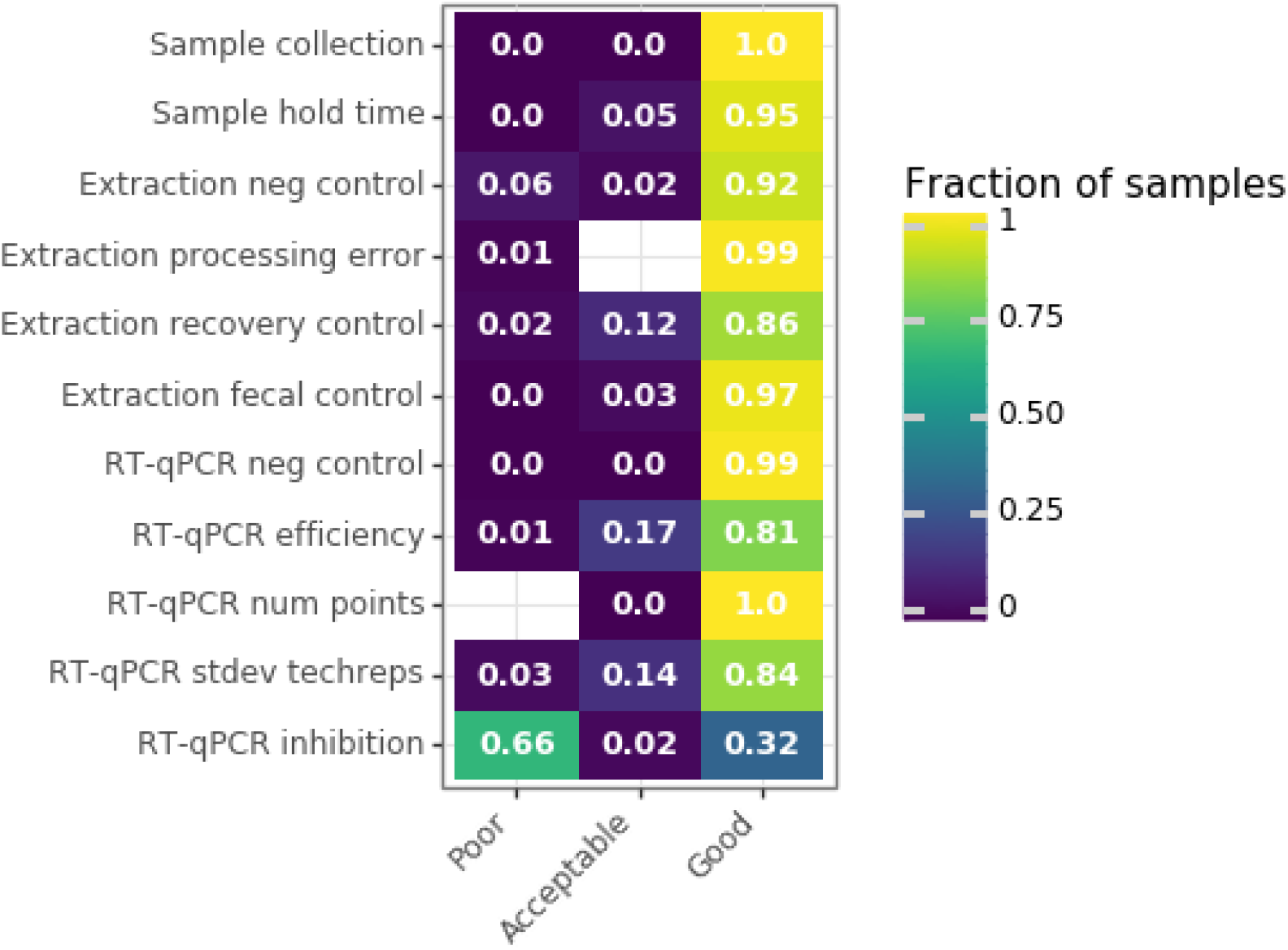
Meta-analysis of quality scores across samples processed in the regional monitoring laboratory. The color and text indicate the fraction of samples that received poor, acceptable, and good scores for each of the eleven quality score parameters. Because missing data can also affect the score, this analysis included only the samples for which N1, PMMoV, and BCoV were assayed and dilutions were performed to assess inhibition in N1 RT-qPCR. Samples (n=1015, including samples that were biological replicates) were taken from residential campuses, sub-sewersheds, and wastewater treatment plant influent.

RT-qPCR inhibition was initially evaluated with a foreign spike-in RNA (Xeno VetMax, ThermoFisher), but the spike-in control was discontinued when results were found to diverge from results of dilution-based inhibition testing in the SARS-CoV-2 N1 assay (**Figure S10**). After mid-November, 2020, inhibition was accounted for by running the N1 assay with undiluted and five-fold diluted RNA template for every sample. While this dilution factor may not completely remove inhibition, further dilution could lead to significant loss of signal. For each sample, results of undiluted and diluted RT-qPCR reactions were compared (**Figure S7**) and inhibition was scored (**Table S4**). Samples that were below the detection limit for N1 in either template dilution were marked “poor” quality for RT-qPCR inhibition. Here, inhibition could not be determined or improved by dilution, increasing the risk of a false negative (**Figure 3**). This included all non-detects, which occurred frequently for residential facilities.

## 4. Discussion

Because the COVID-19 pandemic has spurred widespread application of WBE, we expect that new laboratories will continue to be launched and expanded even after the COVID-19 pandemic. Wastewater monitoring for SARS-CoV-2 requires methods and equipment that may differ from testing wastewater for regulatory compliance and from clinical testing for SARS-CoV-2. In our experience, laboratory set-up required unique considerations for safety, space, workflow, and data management. Additionally, consideration for the scale of the laboratory became important as monitoring needs grew. If warranted, higher throughput than that described here could be achieved via large-volume liquid handling robots for extraction and precision liquid handlers for PCR plate set-up.

Several factors influence whether it is most cost-effective and reliable to test samples in smaller, local laboratories versus larger, centralized laboratories. Smaller labs may face higher costs associated with low-throughput analysis, and potential challenges could arise in data comparison among locations. For larger labs, initial costs of robotic equipment and farther shipping with associated longer turnaround times may be issues of concern. Creating a robust network of WBE monitoring laboratories necessitates capacity-building across sectors including public health labs, utility labs, and commercial labs both nationally and internationally.

Building a flexible data analysis pipeline allowed us to reproducibly compare data over time and to incorporate elements like LoD substitution for non-detects and calculation of geometric standard error of technical replicates. Analysis pipelines like this one can streamline the submission process to CDC-NWSS, which requires extensive details about each sample. Additionally, the pipeline presented here allowed meta-analysis of the data, revealing the variability present in each step of sample processing. While some amount of variation is due to lower quality or technical error, some appears to be inherent methodological variation or variation in the wastewater itself^27,28^. In the future, modeling the contributions of these variables to overall noise in the data could provide better confidence in the results.

The development of a robust quality assurance plan is critical for any laboratory monitoring SARS-CoV-2 in wastewater. Others have documented the controls and standards required for WBE^25,28^. Our newly developed quality scoring framework and code represent one possible model for communicating data quality to partners in public health. Quality score parameters, thresholds, and weights were chosen based on our experience and meta-analyses of our data. While these choices are dependent on the methodology used by a lab, the scoring framework itself is adaptable.

As wastewater monitoring for SARS-CoV-2 continues to expand as part of managing the COVID-19 pandemic, more academic institutions, wastewater agencies, and public health agencies may choose to analyze wastewater samples from routine monitoring efforts in-house. We hope lessons learned and reported here from our experience of developing a laboratory to routinely monitor wastewater samples for SARS-CoV-2 RNA during the COVID-19 pandemic can assist others with similar goals.

## Data Availability

Extended data are available upon request.

## Acknowledgements

The authors gratefully acknowledge the Catena Foundation, the Center for Information Technology in Service to Society, and the Innovative Genomics Institute for funding the UC Berkeley wastewater monitoring laboratory.

## Supporting information captions

SI Table 1. Equipment list for laboratory set-up for the regional wastewater monitoring laboratory at UC Berkeley

SI Table 2. Process flow information for the regional wastewater monitoring laboratory at UC Berkeley

SI Table 3. Per sample cost of consumables and reagents for the regional wastewater monitoring laboratory at UC Berkeley

SI Table 4. Data quality scoring framework

SI Text 1. Supplementary Methods and Figures

SI Text 2. Example biological use authorization for wastewater RNA extraction and RT-qPCR

SI Text 3. Shipping instructions for wastewater treatment agencies

SI Text 4. Labware cleaning standard operating procedure

SI Text 5. Data tables and fields

SI Text 6. Quality assurance and control plan

## References

1. Medema, G., Heijnen, L., Elsinga, G., Italiaander, R. & Brouwer, A. Presence of SARS-Coronavirus-2 RNA in Sewage and Correlation with Reported COVID-19 Prevalence in the Early Stage of the Epidemic in The Netherlands. Environ. Sci. Technol. Lett. 7, 511–516 (2020).

2. Gonzalez, R. et al. COVID-19 surveillance in Southeastern Virginia using wastewater-based epidemiology. Water Res. 186, 116296 (2020).

3. Prado, T. et al. Wastewater-based epidemiology as a useful tool to track SARS-CoV-2 and support public health policies at municipal level in Brazil. Water Res. 191, 116810 (2021).

4. Randazzo, W., Cuevas-Ferrando, E., Sanjuán, R., Domingo-Calap, P. & Sánchez, G. Metropolitan wastewater analysis for COVID-19 epidemiological surveillance. Int. J. Hyg. Environ. Health 230, 113621 (2020).

5. Wu, F. et al. SARS-CoV-2 Titers in Wastewater Are Higher than Expected from Clinically Confirmed Cases. mSystems 5, (2020).

6. Ahmed, W. et al. Comparison of virus concentration methods for the RT-qPCR-based recovery of murine hepatitis virus, a surrogate for SARS-CoV-2 from untreated wastewater. Sci. Total Environ. 739, 139960 (2020).

7. Whitney, O. N. et al. Sewage, Salt, Silica, and SARS-CoV-2 (4S): An Economical Kit-Free Method for Direct Capture of SARS-CoV-2 RNA from Wastewater. Environ. Sci. Technol. (2021) doi:10.1021/acs.est.0c08129.

8. LaTurner, Z. W. et al. Evaluating recovery, cost, and throughput of different concentration methods for SARS-CoV-2 wastewater-based epidemiology. Water Res. 197, 117043 (2021).

9. Philo, S. E. et al. A comparison of SARS-CoV-2 wastewater concentration methods for environmental surveillance. Sci. Total Environ. 760, 144215 (2021).

10. Gerrity, D., Papp, K., Stoker, M., Sims, A. & Frehner, W. Early-pandemic wastewater surveillance of SARS-CoV-2 in Southern Nevada: Methodology, occurrence, and incidence/prevalence considerations. Water Res. X 10, 100086 (2021).

11. Sherchan, S. P. et al. First detection of SARS-CoV-2 RNA in wastewater in North America: A study in Louisiana, USA. Sci. Total Environ. 743, 140621 (2020).

12. Canadian COVID-19 Wastewater Coalition. Phase 1 Inter-Laboratory Study: Comparison of Approaches to Quantify SARS-CoV-2 RNA in Wastewater. https://cwn-rce.ca/report/phase-1-inter-laboratory-study-comparison-of-approaches-to-quantify-sars-cov-2-rna-in-wastewater/ (2020).

13. Pecson, B. M. et al. Reproducibility and sensitivity of 36 methods to quantify the SARS-CoV-2 genetic signal in raw wastewater: findings from an interlaboratory methods evaluation in the U.S. Environ. Sci. Water Res. Technol. 10.1039.D0EW00946F (2021) doi:10.1039/D0EW00946F.

14. Graham, K. E. et al. SARS-CoV-2 RNA in Wastewater Settled Solids Is Associated with COVID-19 Cases in a Large Urban Sewershed. Env. Sci Technol 55, 488–498 (2021).

15. Bivins, A. et al. Wastewater-Based Epidemiology: Global Collaborative to Maximize Contributions in the Fight Against COVID-19. Environ. Sci. Technol. 54, 7754–7757 (2020).

16. Naughton, C. C. et al. Show us the Data: Global COVID-19 Wastewater Monitoring Efforts, Equity, and Gaps. medRxiv 2021.03.14.21253564 (2021) doi:10.1101/2021.03.14.21253564.

17. CDC. National Wastewater Surveillance System. Centers for Disease Control and Prevention https://www.cdc.gov/coronavirus/2019-ncov/cases-updates/wastewater-surveillance.html (2021).

18. Ahmed, W. et al. Surveillance of SARS-CoV-2 RNA in wastewater: Methods optimisation and quality control are crucial for generating reliable public health information. Curr. Opin. Environ. Sci. Health (2020) doi:10.1016/j.coesh.2020.09.003.

19. Whitney, O. Direct wastewater RNA capture and purification via the Sewage, Salt, Silica and SARS-CoV-2 (4S) method. (2020) doi:10.17504/protocols.io.bjr9km96.

20. Greenwald, H. One-Step RT-qPCR for SARS-CoV-2 Wastewater Surveillance: N1, PMMoV, BCoV, SOC. (2020) doi:10.17504/protocols.io.bpk3mkyn.

21. CDC. Wastewater Surveillance Testing Methods. Centers for Disease Control and Prevention https://www.cdc.gov/coronavirus/2019-ncov/cases-updates/wastewater-surveillance/testing-methods.html/#biosafety (2020).

22. FedEx. Packaging UN 3373 Shipments. https://www.fedex.com/content/dam/fedex/us-united-states/services/UN3373_fxcom.pdf.

23. US Department Of Transportation. UN3373 COVID 19 Safety Advisory.pdf. https://www.phmsa.dot.gov/sites/phmsa.dot.gov/files/2020-06/UN3373%20COVID%2019%20Safety%20Advisory.pdf.

24. University of California, Berkeley, Environmental Health & Safety. Hazardous Waste Management. http://ehs.berkeley.edu/sites/default/files/lines-of-services/hazardous-materials/52hazardouswaste.pdf (2014).

25. Ahmed, W. et al. Surveillance of SARS-CoV-2 RNA in wastewater: Methods optimisation and quality control are crucial for generating reliable public health information. Curr. Opin. Environ. Sci. Health (2020) doi:10.1016/j.coesh.2020.09.003.

26. Burns, M. J., Nixon, G. J., Foy, C. A. & Harris, N. Standardisation of data from real-time quantitative PCR methods – evaluation of outliers and comparison of calibration curves. BMC Biotechnol. 5, 31 (2005).

27. Feng, S. et al. Evaluation of sampling frequency and normalization of SARS-CoV-2 wastewater concentrations for capturing COVID-19 burdens in the community. medRxiv 2021.02.17.21251867 (2021) doi:10.1101/2021.02.17.21251867.

28. Ahmed, W. et al. Minimizing Errors in RT-PCR Detection and Quantification of SARS-CoV-2 RNA for Wastewater Surveillance. (2021) doi:10.20944/preprints202104.0481.v1.

