## Supplementary Information for "Operationalizing a routine wastewater monitoring laboratory for SARS-CoV-2"

**Supplementary Methods**

The data analysis pipeline was written to convert Cq values to gene copies per well using the slope and intercept from the standard curve on each plate. However, we found that the intercept of the standard curve shifted more than expected from plate to plate (**Figure S9**). We attribute this to differences in the volume of standard pipetted into the top well of the serial dilution by multiple technicians. To minimize variation in the data due to the standard curve, the data analysis pipeline was used to apply a single average standard curve for SARS-CoV-2 N1 calculated from 145 individual plates. Other sources of plate-to-plate variability were limited by keeping constant the mastermix, plate type, qPCR machine, and primer/probe mixture. To improve consistency of standard curves, a single technician dilutes and aliquots all points on the curve weekly for each assay. This change has substantially improved the consistency of the intercepts across plates.

**Supplementary Figures**

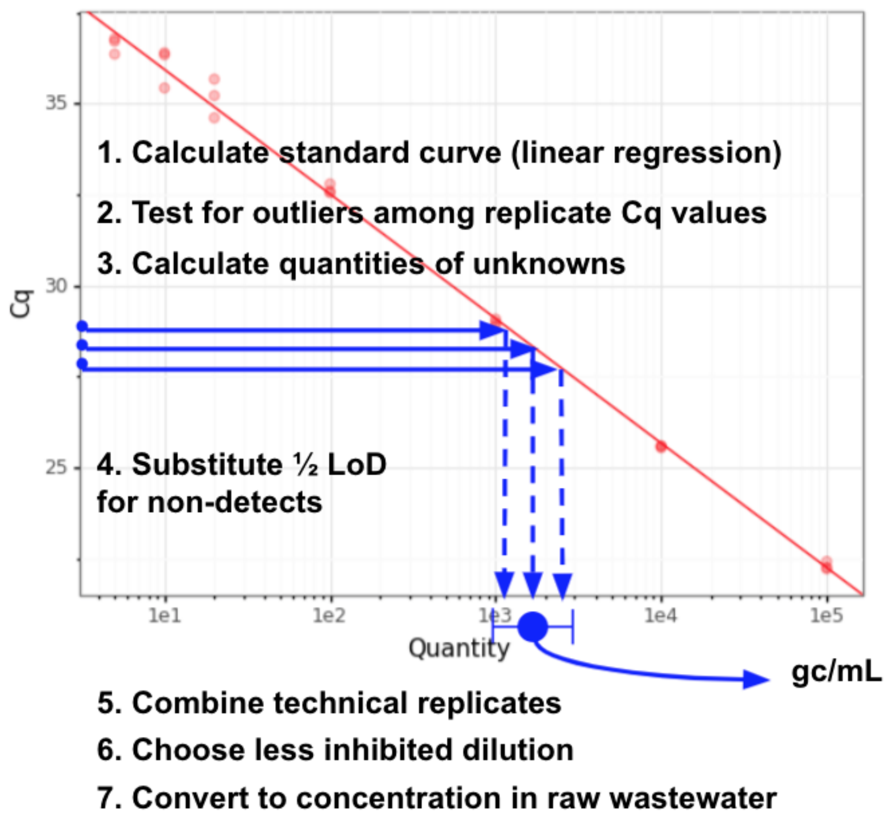

**Figure S1.** Schematic of data analysis pipeline. For more details see README and code at https://github.com/wastewaterlab/data_analysis.

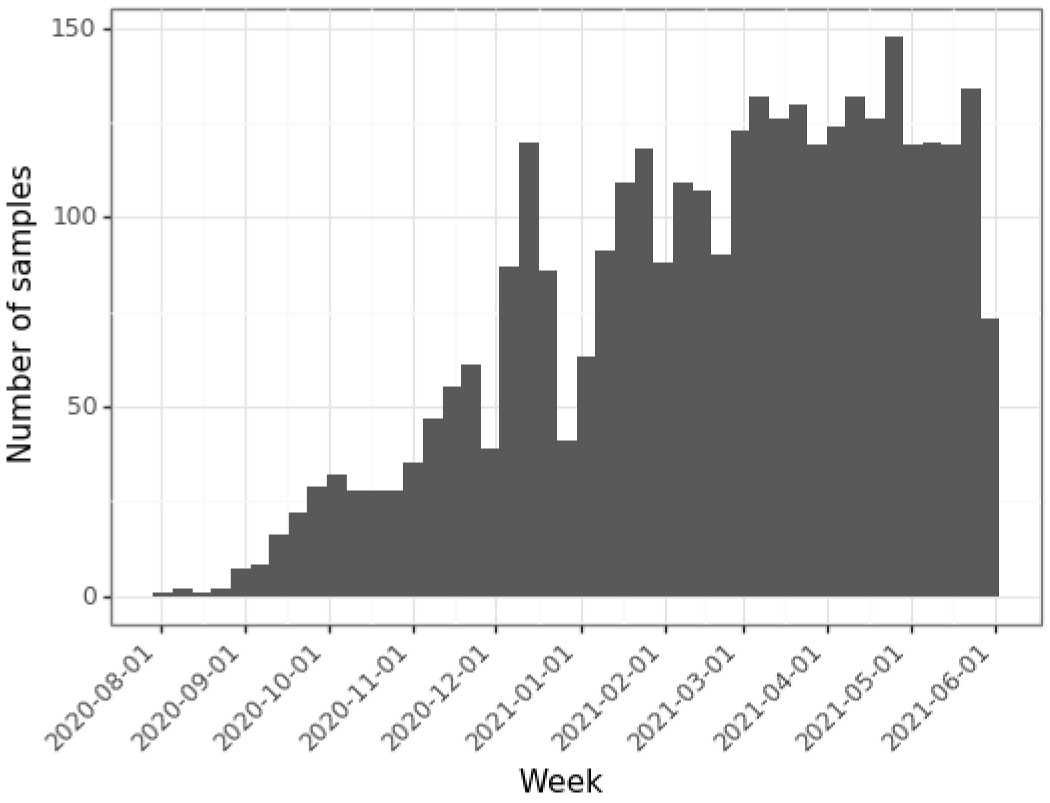

**Figure S2.** Weekly sample counts over time. Counts include biological replicates processed in parallel.

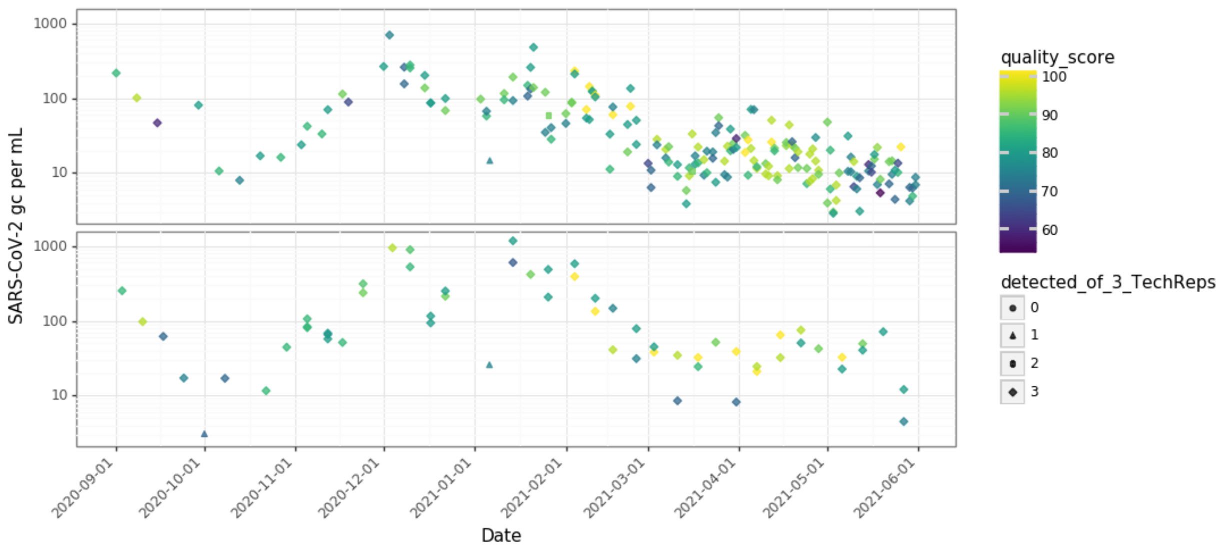

**Figure S3.** Scored data for biological replicates as shown in Figure 2.

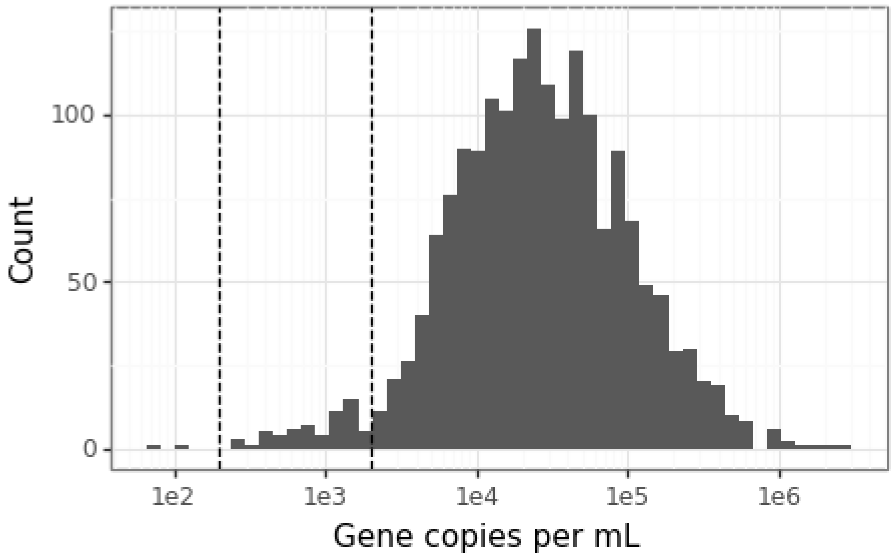

**Figure S4.** Log10 distribution of PMMoV concentrations across samples. Dashed lines are shown at 200 and 2000, indicating chosen thresholds for acceptable and good data.

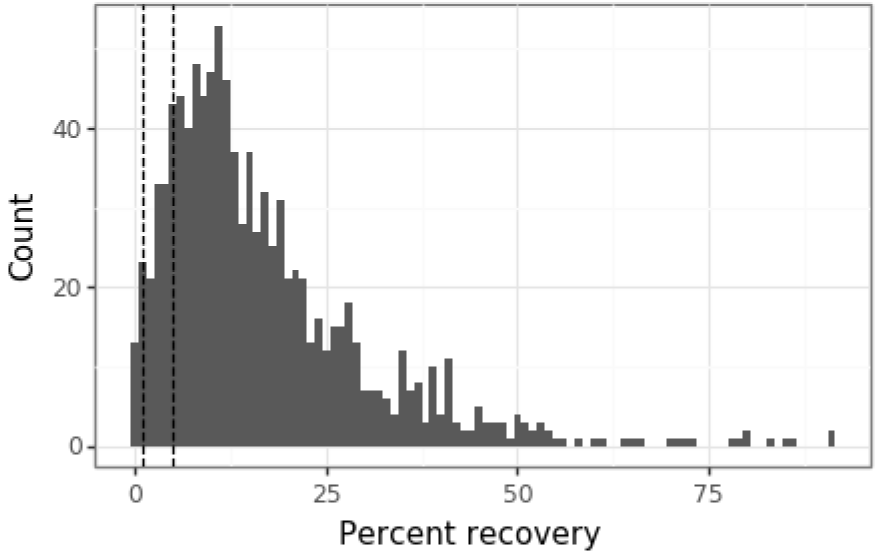

**Figure S5.** Distribution of recovery efficiency of bovine coronavirus spike-in control across samples. Dashed lines are shown at 1% and 5% recovery, indicating chosen thresholds for acceptable and good data.

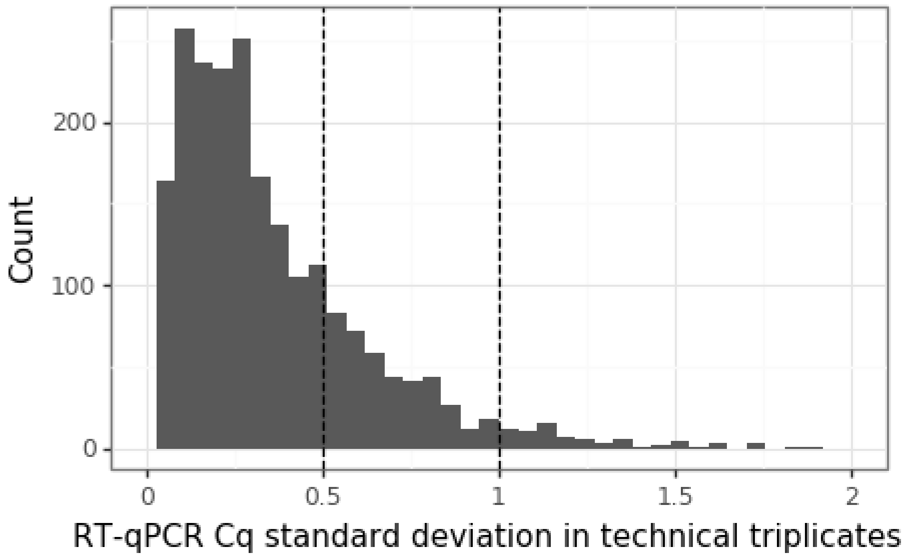

**Figure S6.** Distribution of standard deviation for Cq values in triplicate qPCR wells across samples. Dashed lines are shown at 0.5 Cq and 1 Cq, indicating chosen thresholds for acceptable and poor data. Data are shown only for samples in which all three technical replicates amplified. Two samples had Cq standard deviations greater than 2 and are not shown.

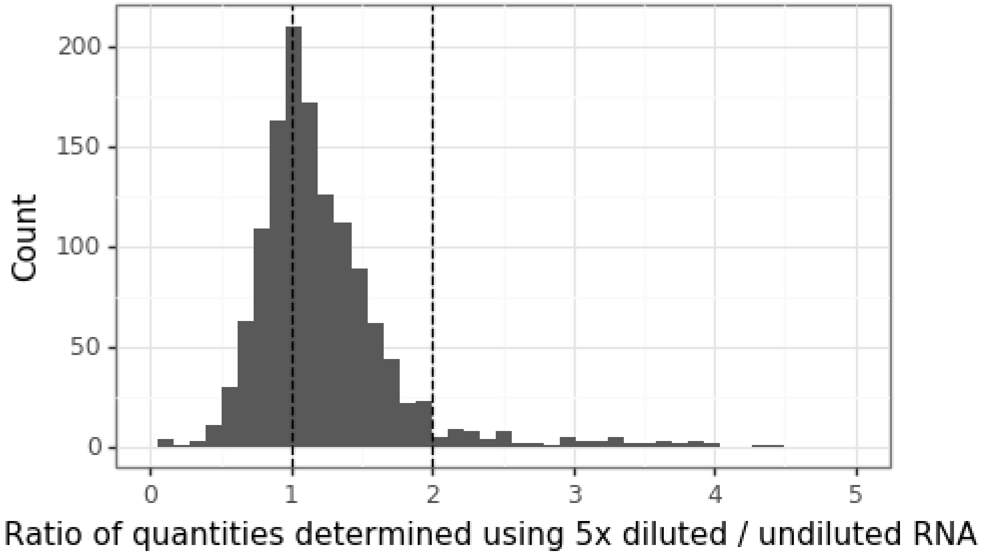

**Figure S7**. RT-qPCR inhibition varied across samples. The CDC N1 assay was performed on undiluted template and 5-fold diluted template (in triplicate for each dilution). The average gene copies per well were calculated for each dilution and multiplied by the dilution factor. For samples in which both dilutions were above the detection limit (n = 1335), dilutions were compared to each other to determine the severity of inhibition. The line at 1 indicates no inhibition, while samples to the right of this line were potentially inhibited, with some variability due to pipetting error. Samples that were inhibited above a 5x-to-undiluted ratio of 2 were scored as “poor” in the quality score. There were 33 inhibited samples with ratios greater than 5 that are not shown.

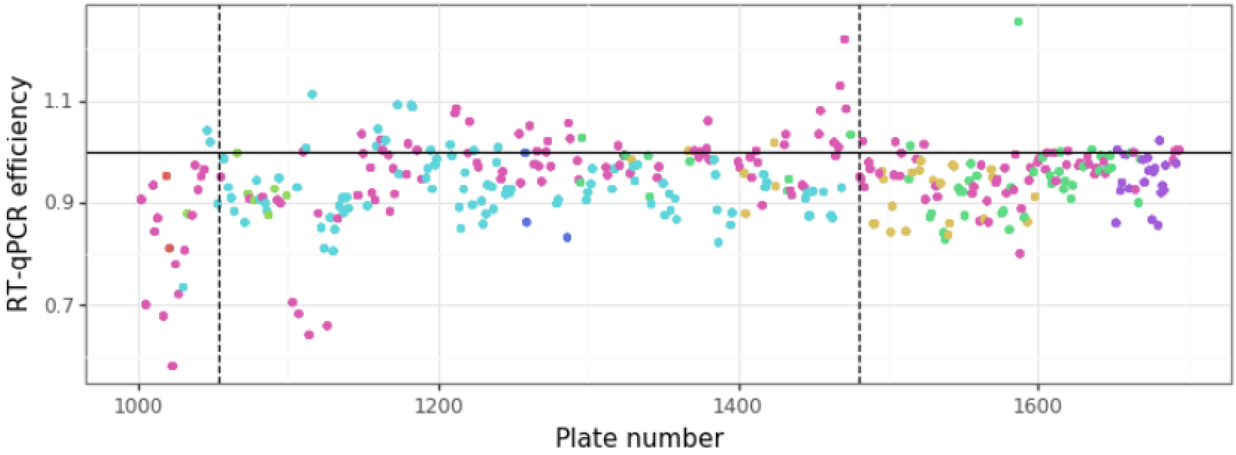

**Figure S8.** RT-qPCR efficiency varied by technician (color) and over time. Plates plotted to the left of the dashed line at 1054 used RNA standards and experienced high ambient temperatures. Subsequent plates used DNA standards and were assembled in a temperature-controlled room. Plates to the right of the dashed line at 1480 were made using pre-aliquoted standard curves prepared weekly (see supplementary methods above).

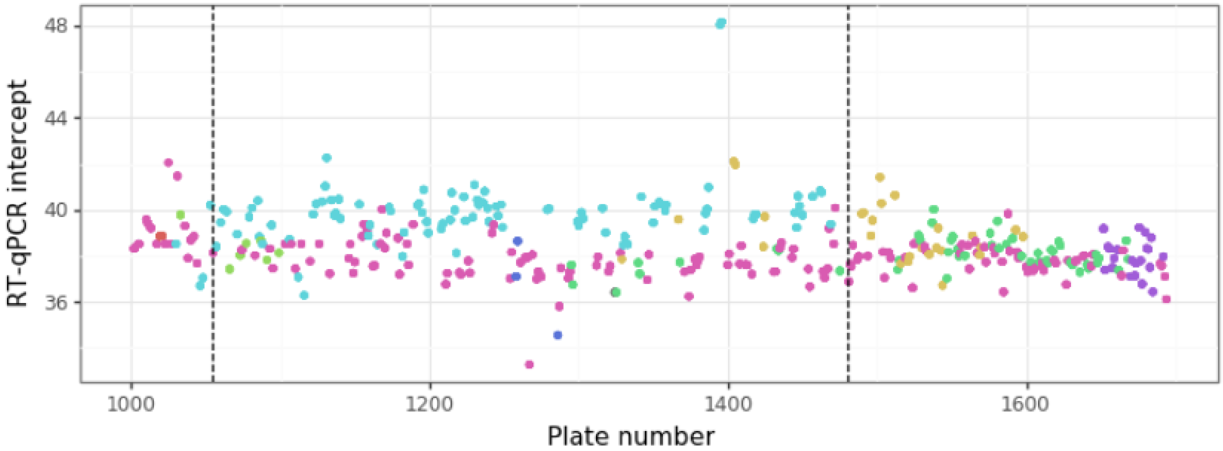

**Figure S9.** RT-qPCR intercept varied by technician (color) and over time. Plates plotted to the left of the dashed line at 1054 used RNA standards and experienced high ambient temperatures. Subsequent plates used DNA standards and were assembled in a temperature-controlled room. Plates to the right of the dashed line at 1480 were made using pre-aliquoted standard curves prepared weekly (see supplementary methods above).

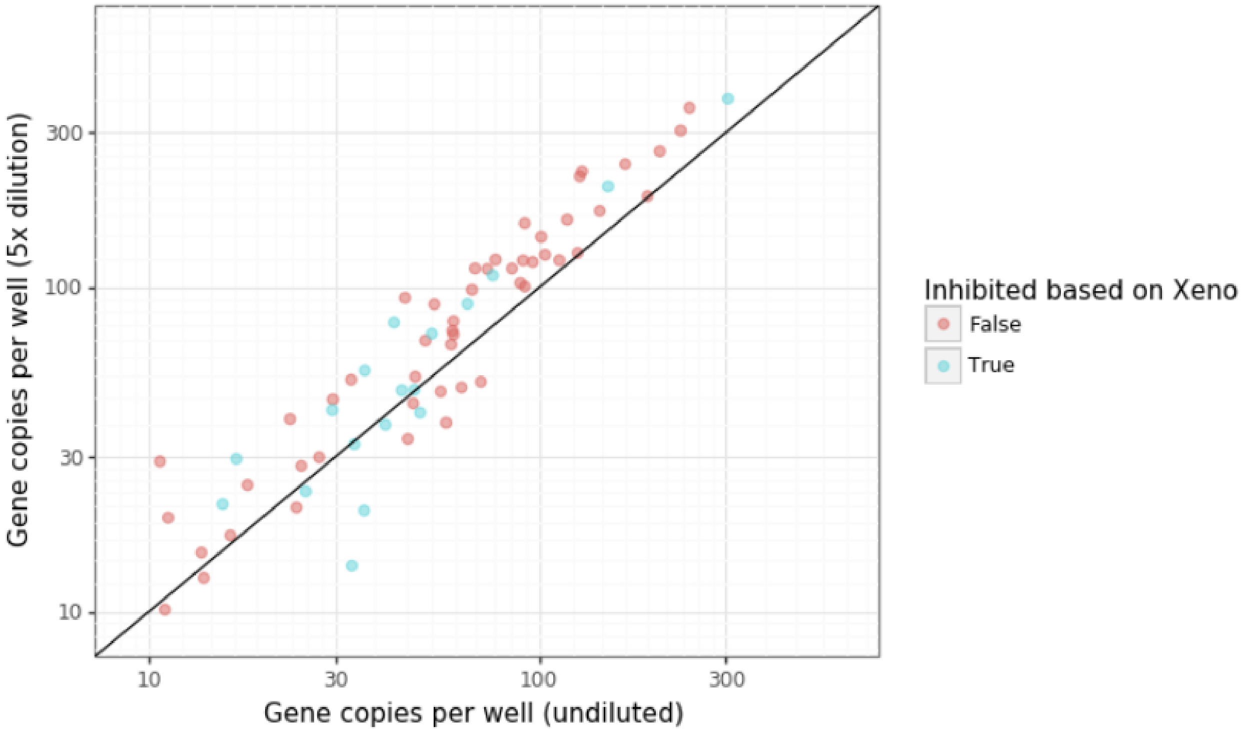

**Figure S10.** Comparison of spike-in RNA control and dilution methods for inhibition testing. Each point represents a single sample for which SARS-CoV-2 N1 was quantified as gene-copies per well using undiluted (x-axis) and 5-fold diluted (y-axis) template. Gene copies (log10-scale x- and y-axes) are adjusted by the dilution factor. N1 was multiplexed with the Xeno VetMax assay by ThermoFisher. Blue points indicate that VetMAX Xeno was > 1 Ct higher in the sample than in the water control on the same plate, suggesting the sample was inhibited. Points for samples without inhibition can be expected to appear near or below the 1:1 line, whereas points for samples with inhibition will have higher effective concentrations after dilution of inhibitors. If the dilution method and Xeno method agree, blue points should be consistently above the 1:1 line, while red points should be consistently near or below the line (assuming some pipetting error in dilutions and RT-qPCR).

#### Example biological use authorization for wastewater RNA extraction and RT-qPCR

##### Do Not Fill Table-Completed by the Biosafety Officer

| BUA Number | Level of Containment | DATE of Approval | CLEB Representative |
|---|---|---|---|

BUA No. __ Expiration date: ____________
Name of Principal Investigator: ___________
Title: ________ Phone Number: _________
Department: CEE Mail Code: _____
E-mail Address: _____________ FAX Number: _____
Funding Source(s): _____ Grant Number(s): _____

The purpose of this form is to record any changes to a previously approved Biological Use Authorization. The primary use of this form is to amend any BUA for the use of recombinant DNA molecules. The National Institutes of Health (NIH) requires UC Berkeley to review all recombinant DNA experiments. Any changes to experiments involving the generation or use of recombinant DNA molecules conducted under **Sections III-A-1 through Section III-F-6 of the NIH Recombinant DNA Guidelines** must be approved prior to the initiation of the changes involving the scope of work. In addition, any changes to work involving Risk Group 1through Risk Group 3 infectious agents, exclusive of recombinant DNA also must be reflected on a completed BUA Amendment Form. If you have any questions, please call 643-6562.

##### Requested Modifications-Explain any changes in detail on page 3

##### 1. Changes in Location of experiments:

| ACTION | | | BUILDING | ROOM | BIOSAFETY LEVEL (BL-1, BL-2) | SHARED ROOM* |
|---|---|---|---|---|---|---|
| Add | Delete | Modify | | | | |
| ☐ | ☐ | ☐ | XXX Hall | 101 | BL-2 (used to be BSL-3) | ☐ Yes |
| ☐ | ☐ | ☐ | | | | ☐ Yes |
| ☐ | ☐ | ☐ | | | | ☐ Yes |

##### 2. Changes in Personnel:

| ACTION | | NAME | | Position | Agents and materials | e-mail | Training date | |
|---|---|---|---|---|---|---|---|---|
| Add | Delete | Last | First | | | | Bio safety | BBP * |
| ☐ | ☐ | Snow | John | Graduate Student Researcher | *wastewater* | | completed | |
| ☐ | ☐ | | | | | | | |
| ☐ | ☐ | | | | | | | |
| ☐ | ☐ | | | | | | | |

* Bloodborne Pathogen (BBP) training is required for all who work with lentiviral vectors human material or cell lines, non-human primate samples or cell lines. Please use https://jwas.ehs.berkeley.edu/lab/ for training reference – and either state "up-to-date", or the date training will be completed.

##### 3. Changes to Biological Agents/Toxins used in experiments:

Addition of Agents/Toxins: ☐ Yes ☒ No
Deletion of Agents/Toxins: ☐ Yes ☒ No
Modification of use: ☐ Yes ☐ No
Addition of Human Blood, blood product, unfixed tissue, cell culture ☐ Yes ☒ No
Addition of experiments involving animals ☐ Yes ☒ No **(if yes, complete item 5)**

**Complete all applicable fields in the table:**

| Agent | Pathogen | | | Select Agent | | Risk Group* | **Used in** | | **Toxin** | |
|---|---|---|---|---|---|---|---|---|---|---|
| | Human | Animal | Plant | CDC | USDA | | **TC** | **Animal-specie** | **Yes** | **$LD_{50}$ or LDL** |
| *wastewater** | ☐ | ☐ | ☐ | ☐ | ☐ | RG2 | | | ☐ | |
| | ☐ | ☐ | ☐ | ☐ | ☐ | | | | ☐ | |
| | ☐ | ☐ | ☐ | ☐ | ☐ | | | | ☐ | |
| | ☐ | ☐ | ☐ | ☐ | ☐ | | | | ☐ | |

*Wastewater is currently elevated to BSL-2+ because of the added unknown risk of SARS-CoV-2 within and our protocols reflect this with added safety measures.

#### 4. Recombinant DNA experiments:

Vector currently used:
☐ adenovirus
☐ retrovirus ☐ amphotrophic ☐ ecotropic
☐ lentivirus
☐ other:

*Will transgene increase the host range of disease to humans ☐ Yes ☐ No

**For recombinant DNA experiments, complete the outline:**
**Host**:
**Vector**:
**Nature of inserted sequences**:
**Source of inserted sequences**:
**Types of manipulation:**
**Attempt to express foreign gene:**
**Protein produced:**
**Containment/Biosafety Level:** ***BSL*________**
**Section of Guidelines:**

#### 5. Description of Animal Experiments

**Species of animal used: _____**
**Potential Risks with Agent Use**

**A. Release or Shedding through:**
☐ Feces/Urine
☐ Bloodborne
☐ Respiratory Secretion
☐ Fomite/Cutaneous
☐ Other: _____

**B. Physical Risks**
☐ Sharps/Laceration
☐ Ocular
☐ Bite/Scratch
☐ Respiratory/Allergen
☐ Other: _____

**C Location of:**
**Animal Procedures:**
**Animal Housing:**
**Storage freezer:**
**Carcass disposal:**

**D. Transportation**
**Describe method of transport of agents and/or animal between multiple locations, if applicable:**
Wastewater samples will be transported to the lab in personal vehicles or through FedEx mail.
For samples that will be transported to the lab from sampling sites in personal vehicles, wastewater samples will be transported to the lab in tertiary containment (labeled primary container, secondary bag or container, and a cooler) in personal vehicles by John Snow. Samples will then be processed following the protocol below or stored in secondary containment in a designated -80 C freezer or 4 C fridge in the BSL2+ lab.
For samples that are shipped from wastewater treatment facilities to the lab, three 50 mL plastic centrifuge tubes filled with NaCl, Tris, and EDTA will be sent to the treatment facility in advance of sample collection. Wastewater agency staff will pour 40 mL of raw wastewater into each tube, close the tube, wipe it with a disinfecting wipe, then tape the lid shut. They will then put the tubes in a Ziploc bag with absorbent material. There is sufficient absorbent material to contain the sample in the unlikely event of a complete spill. The bag with the tubes will be placed in the box, followed by an ice pack. The box will then be sealed with the biohazard label (UN3733) affixed and shipped via FedEx Overnight. All supplies and step-by-step instructions will be sent to the wastewater treatment facility.

6. **Summary Statement of Changes: Please write a paragraph that will describe the procedures used and the potential implications for health and safety.**

**Summary of Experimental Goals:**
Our research is scaling up to receive wastewater samples from around the San Francisco Bay Area for regional monitoring of wastewater for SARS-CoV-2 and requires additional lab space. Approximately XXX samples will be processed per day in the lab. Samples will either be dropped off to our lab by wastewater agency employees, picked up by lab members in personal vehicles, or shipped to our lab via FedEx following the previously described protocols (see Section 5.D. Transportation).

Briefly, this research involves collection, RNA extraction, and RT-qPCR of wastewater samples for the detection and monitoring of SARS-CoV-2 in the San Francisco Bay Area (with possibility to expand to a wider region). This research will help communities track the SARS-CoV-2 prevalence in their wastewater in order to allocate medical supplies or alter public health measures. Samples provided to us will be both from the headworks of municipal wastewater treatment facilities as well as from locations higher up in the sewershed, including areas drawing from the UC Berkeley campus. In general, wastewater treatment plant operators (or EH&S employees at UC Berkeley sampling sites) will use ISCO composite samplers to collect the wastewater from manholes or other centralized collection points. The samples provided to researchers will be in 40 mL volumes, already prepared in tertiary containment.

**Lab space:**
We will conduct work with samples in the BSL-2 approved space—previously BSL-3—in XXX Hall. There are 2 emergency eyewash stations, 1 shower, and an emergency exit. The lab space is comprised of multiple connected sub-rooms, and the lab activity that will be conducted in each is described below:

- Room 101: Entryway. No samples will be handled here outside of tertiary containment. Generally, only used as a passageway, with some storage and trash bins. In this room there is no sample of bio waste because samples will all be processed further in the lab. No lab work will be conducted in this space. BSL-1.
- Room 101AA: Shipping and storage room. No samples will be handled here outside of tertiary containment. 50 ml screw top conical tubes will be filled with NaCl to be sent out for wastewater fulfillment at site. These will be placed in Ziploc bags which in turn will be placed in cardboard boxes which will be labeled with FedEx labels with corresponding addresses. No lab work will be conducted in this space. BSL-1.
- Room 101B: Donning/doffing room including storage lockers for personal belongings, handwashing sink, and emergency shower. No samples will pass through this room outside tertiary containment. No lab work will be done in this space. BSL-1.
- Room 101A: Autoclave receiving room. Clean items will be removed from the autoclave. BSL-1. No lab work will be conducted here.
- Room 101BA: Sample receiving and buffer preparation room. This room will be used to weigh out salts and make buffers. No samples will be handled outside of secondary containment in this room. BSL-1. Only preparation will be done in this space, no processing of samples.
- Room 101C: Main laboratory room. Samples will be unboxed (taken out of tertiary containment, remaining in secondary containment) in this room before being placed in the refrigerator in secondary containment. This room will contain 3 functioning, certified class II biosafety cabinets in which samples will be processed. The biosafety cabinets will be in the corners of the room such that researchers will always be greater than 6 feet apart. The refrigerator and freezers for sample storage will also be located in this room. This room will also have qPCR instruments, storage, and computer workstation(s). This room is the only room with a lab sink which will mainly be used for dishwashing. Anyone working in this room must be listed on the BUA. This room will follow BSL-2+ safety precautions.

- Room 101CA: A liquid-handling robot for routine qPCR set-up will be housed in this room. N95 masks will be required for working in this room due to the possibility of exposure to aerosolized SARS-CoV-2 RNA. This room will follow BSL-2+ safety precautions.

**Protocols:**
We will conduct all work with samples in the class II biosafety cabinet in the BSL-2 approved space in XXX Hall room 101 with the door closed. Sample containers will be opened only inside a BSL-2 Class II type A2 biosafety cabinet. Researchers will utilize full BSL-2+ PPE including laboratory coats, splash goggles, and double gloves while working with biological samples in the biosafety cabinet at all stages.

We follow additional safety precautions beyond typical BSL-2 measures based on CDC guidelines for working with environmental samples during the COVID-19 pandemic. Researchers will don and doff PPE in a designated location by the entrance next to the handwashing sink. Any activity in which the wastewater will be removed from secondary containment will be conducted in a biosafety cabinet. For any step involving vortexing, samples will be allowed to sit before containers are opened for as long as the protocol allows. After 40 mL wastewater samples are spiked with bovine coronavirus and vortexed, they will sit in a dry bath 70 C for 45 minutes, allowing time for aerosols to settle before containers are opened.

The 4S method for direct extraction from wastewater will be employed (see below for method protocol). Briefly, high concentrations of salt are added to lyse viral particles in the sample and inactivate RNAses. The samples are heat-inactivated (at 70 C for 45 minutes including a temperature control tube to ensure proper inactivation), filtered, ethanol precipitated, passed through a silica column, then eluted. Any activity in which the wastewater will be removed from secondary containment will be conducted in a biosafety cabinet, including steps after heat inactivation.

This lab space will also follow qPCR set-up protocols either by hand or in the automated liquid-handler. When qPCR work is set up by hand, all work will be conducted in a biosafety cabinet. Set-up for qPCR conducted by the liquid-handler is the only lab work that will be conducted outside the biosafety cabinets. The liquid-handler is contained in a polycarbonate shell, but we wanted to take additional safety precautions. Due to the liquid-handler not being completely contained, all users of the liquid-handler and those working in the room while the liquid-handler is operational will wear N95 masks. Additionally the liquid-handler is located in the room with the lowest pressure, thus reducing the spread of aerosols to other rooms.

**Lab Safety:**
NOTE: To date, no infective SARS-CoV-2 has been found in wastewater. Rimoldi et. al (2020) added concentrated wastewater to VERO E6 cells and did not observe any cytopathic effect. (https://www.sciencedirect.com/science/article/pii/S0048969720344405?via%3Dihub).

In the event of a spill, the area will be wiped down and disinfected with 10% bleach for 20 minutes. Spills would likely be contained within the biosafety cabinet. Maximum volume handled is 40 mL. Spills will be reported to EHS ( and/or XXX-XXX-XXX) within 2 hours. BSL-2 waste will be disposed of in red biohazard waste bags, and pickup will be requested within 1 week. To reduce aerosol risk, we will keep centrifuged samples firmly capped during centrifugation, and vortexing and pipetting will be performed slowly and in the biosafety cabinet. Excess wastewater samples will be disinfected for 20 minutes by adding bleach to a final concentration of 10% before drain disposal. Sample tubes, filters, and pipette tips in contact with samples will be disposed of as biohazardous waste (red biological waste bags), and pickup will be requested within 1 week of waste placement in the bin.

Disinfectants: 10% chlorine bleach and 70% ethanol will be used as disinfectants for work surfaces and sampling instruments. Bleach and ethanol will be in labeled, unbreakable containers in 500mL quantities or less. 10% bleach will be renewed every two weeks.

Our team will coordinate carefully to follow guidelines on social distancing. Each team member has applied for authorization to be on campus. We will limit the workers in the lab space to maintain social distancing, and all individuals will be wearing BSL-2 PPE (when they are handling samples) or a cloth or surgical mask (when not handling samples) or an N95 mask (when interacting with the liquid-handler). Each team member will perform hand sanitizing and hand washing when leaving the lab. Team members will be briefed on the importance of self-observation for COVID-19 symptoms. If a team member reports fever, cough, or other potentially infectious symptoms, they will stay at home, self-isolate for at least 14 days from symptoms, and inform a doctor. EH&S will also be notified about any team member with symptoms that may be related to laboratory exposure.

**Extraction method: Direct RNA extraction via Sewage, Salt, Silica and SARS-CoV-2 (4S): A rapid, economical, and kit-free method for direct-capture of wastewater RNA**
This rapid and economical method employs common laboratory reagents to extract and directly capture RNA from wastewater using salt-based lysis and silica mediated nucleic acid capture to recover high quality wastewater RNA.
Sample volumes of 40 mL have proven sufficient to detect low-abundance viruses such as SARS-CoV-2 present in Bay Area wastewater. Further, the method enriches all wastewater RNAs without bias, enabling the simultaneous purification and detection of viral RNAs of interest alongside wastewater human fecal load markers suitable for “fecal load” normalization. Below is a protocol of the steps needed to extract RNA from 40mL of wastewater.

**Required reagents and equipment:**
Reagents:

- NaCl
- EDTA
- Tris(hydroxymethyl)aminomethane (TRIS), pH 7.2
- Ethanol

Equipment:

- Silica column (Zymo III-P, cat #C1040-5)
- 5uM filter (Durapore SVLP/PVDF)
- Vacuum manifold
- Microcentrifuge capable of 10,000xg speed

**Sample preparation for 40 mL wastewater (Lysis & Filtration)**

1. Add 9.35 grams of NaCl (final concentration 4 M), EDTA to a final concentration of 1 mM and Tris (pH 7.2) to a final concentration of 10mM.
2. Agitate sample until all NaCl dissolves. Vortex or shake for 15 seconds.
3. Heat inactivate at 70 C for 45 minutes in a dry bath or water bath.
4. Filter lysed sample through a 5 µM PVDF filter via vacuum or syringe filtration.
5. Proceed to RNA extraction.

**Direct RNA extraction (RNA Binding, Washing, Eluting)**

1. Add 1 volume (40 mL) of 70% ethanol to lysed and filtered sample.
    1. Agitate sample to mix ethanol and wastewater lysate.
2. Pass the 80 mL lysed and ethanol treated sample through Zymo III-P silica column using a vacuum manifold.
3. Pass 5 mL of 4SWB1 through the column to wash.
4. Pass 10 mL of 4SWB2 through the column to wash.
5. Detach the reservoir from the column and centrifuge the column at 10000xg for 1 minute to remove residual 4SWB2.
6. Add 200 uL of elution buffer (Commercial column elution buffer, e.g. ZymoPURE, or pH 8 TE buffer) preheated to 50 C.
    1. Incubate columns with the elution buffer for 1 minutes.

7. Place columns into LoBind 1.5 mL microfuge tubes, and spin at 10,000xg for 5 minutes to elute bound RNA.
8. The eluted RNA is now present in the spun-through eluate. Proceed with downstream RNA analysis via qRT-PCR, sequencing or other.

**Principal Investigator Signature: ________________ Date: _____________**

RAW WASTEWATER SAMPLING KIT INSTRUCTIONS

**Sampling Kit Contents:**

- Sample collection tubes pre-filled with salt mix (to preserve the viral genetic material in the sample; non-hazardous)
  - 3 labeled sample tubes are included per sampling location
- Ice pack
- Sealable plastic bag with absorbent material
- Return label
- Packaging tape (included in first kit only)

**What you need:**

- A sampling device to collect a 24-hour composite wastewater sample with clean bottles
- Flow rates and other metadata associated with the sample
- Personal protective equipment

**Please plan to ship the samples to the laboratory on the day they were collected. Call FedEx at 1-800-463-3339 to schedule your pickup via overnight shipping.** Samples should be shipped as biohazard class B according to the following guidelines (FedEx guidelines, IATA guidelines, PHMSA guidelines).

**Protocol:**

1. Carefully open your box (you will use the same box to send back the samples).
2. Retrieve the ice pack from the box. Store the ice pack in a freezer.
3. Collect a 24-hour composite sample of sewage from the designated collection point(s).
4. Mix the sewage (invert bottles or stir) then add 40 mL to each of the provided sample collection tubes (three 50 mL tubes, all with salt, found in the sample kit). **After adding 40 mL of sample, the total volume should be at the 45 mL line for the tubes.**
   a. Fill the 3 tubes for each sample location.
   b. Check the sample label to ensure that the tubes being filled have the same label except for the last digit (e.g., B_loc_072120_1, B_loc_072120_2, etc.).
      i. If the date (ddmmyy, in this example 072120) is missing from the label please fill it in.
5. Screw the sample tube caps on tightly. Wrap packaging tape around the cap to create a secondary seal.
6. If you're collecting samples from multiple locations, repeat steps 3, 4, and 5.
7. Invert each sample tube at least 20 times to mix and completely dissolve the salt.
8. Wipe down the outside of the tubes.
9. Place the tubes in the provided plastic bag with the absorbent material. Seal the bag.
10. Put the bag and the frozen ice pack into the shipping box.
11. Fill in the sampling log and metadata chart via the google forms.
12. Close the shipping box and attach the provided shipping label and biohazard label to the outside, on top of the previous shipping label.

13. Ship the samples on the day they were collected. Call FedEx at 1-800-463-3339 to schedule your pickup via overnight shipping.

### Labware Cleaning Standard Operating Procedure

**After RNA extraction the following items need to be cleaned:**

**Filter holders and bottles**

- Unscrew filter holders and remove filters, place filters in biohazard bin
- In an autoclave bin, soak filter holders in 10% bleach solution for 30 mins before removing from biosafety cabinet
- Place filter holders in DI water rinse bath and move around to rinse
- Repeat rinsing steps 3 times
- (autoclaving is optional but decreases the lifespan of the plastic, so we aren't doing it)

**Vacuum manifold**

- Pour out waste into waste bottle
- Transfer liquid waste to the ethanol waste bottle

**Vacuum waste trap**

- Empty vacuum trap waste into ethanol waste bottle

**Biosafety cabinet**

- Spray all items with bleach and 70% ethanol before removing from biosafety cabinet
- Once cabinet is empty, spray surface and walls of biosafety cabinet with 10% bleach and allow to sit
- Wipe down surfaces
- Spray surface and walls of biosafety cabinet with 70% ethanol and wipe clean
- Be sure to remove all bleach to prevent corrosion
- Close sash
- Disinfect with UV light for 15 min
- Turn off lights and blower
- Remake 10% bleach weekly

### Data tables and fields

#### From wastewater agency

**Sample collection log**
- utility_name
- sample_code
- time_of_sample_collection
- date_sampling
- Additional notes about the sample
- Composite sampler log (optional file upload)
- Flow (MGD) for this location, if known
- Sample_type (composite or grab)
- Number of samples composited
- Number of hours represented by composite
- Sample draw frequency (per hour)
- Chain of custody electronic signature

**Sample metadata log**
- utility_name
- sample_code
- date_sampling
- TSS (mg/L) (if available)
- COD (mg/L) (if available)
- BOD (mg/L) (if available)

#### From laboratory

**Facility table**
- utility
- agency
- county
- Interceptors
- Day of Sampling
- Full name
- Primary_contact_name
- Primary_contact_email
- Lab_contact
- Lab_contact_email
- Shipping address

**Shipping table**
- Sampling Week
- utility
- Location
- Interceptors
- Number of Samples
- Tracking number
- Return Tracking number

**Site table**
- utility_name
- County
- Utility
- Facility
- site
- sample_code
- sample_level
- sampling_days
- sampling_frequency_per_week
- site_full_name
- site_description
- site_pretreatment
- site_population_served

**Sample table**
- sample_id
- sample_code
- date_sampling
- replicate
- batch
- date_extract
- extracted_by
- weight
- bCoV_spike_tube
- GFP_spike_tube
- processing_error
- qPCR_batch
- Storage_conditions
- salted_before_freezing
- date_frozen
- elution_vol_ul
- effective_vol_extracted_ml
- weight_vol_extracted_ml
- bCoV_spike_vol_ul
- GFP_spike_vol_ul
- storage_notes

**Sample naming scheme:**

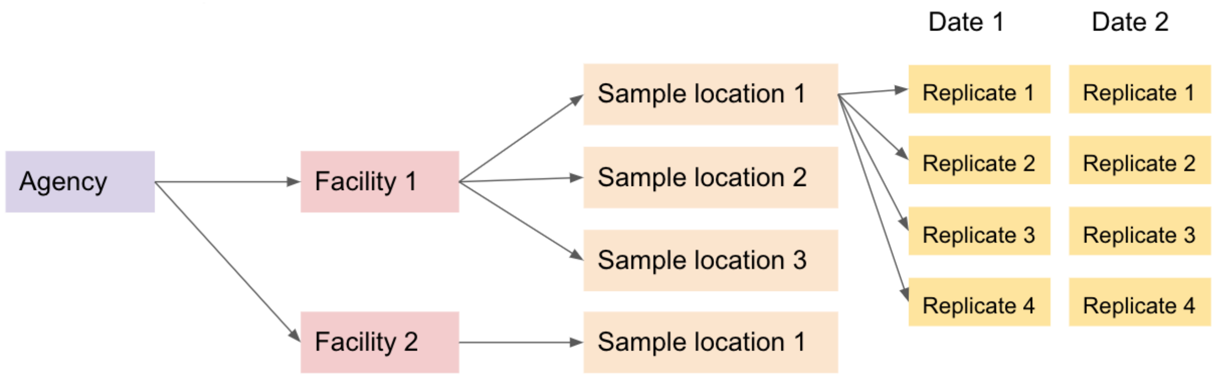

**Sample Collection:**

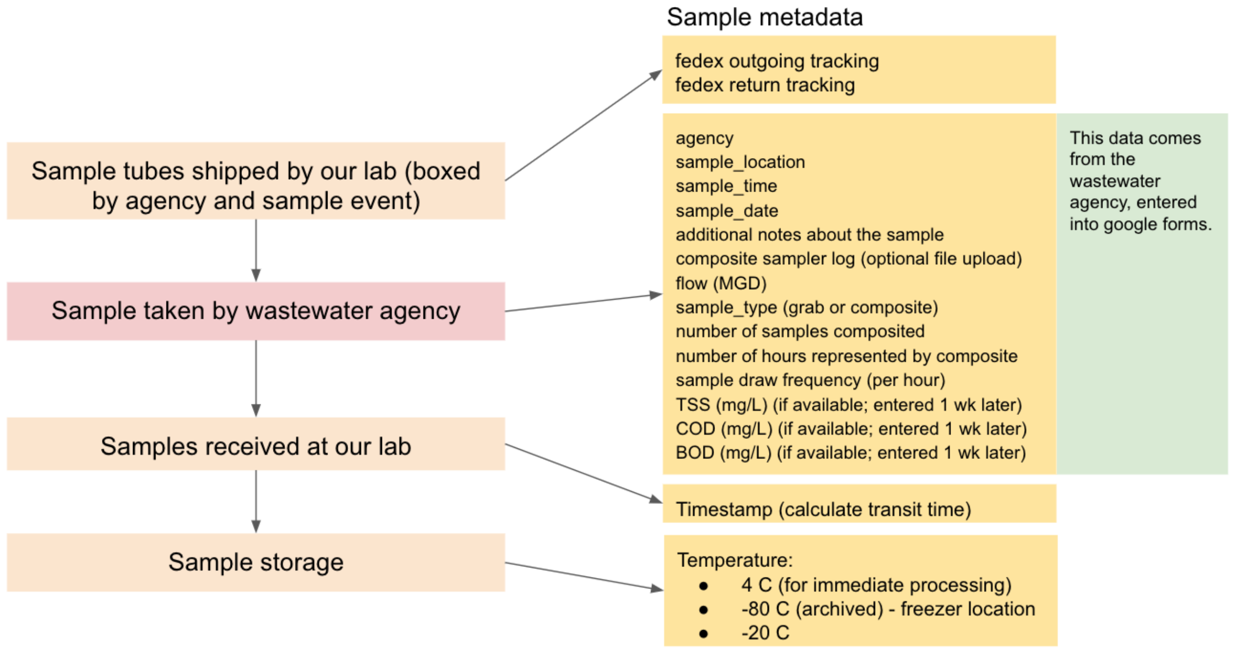

#### Sample processing:

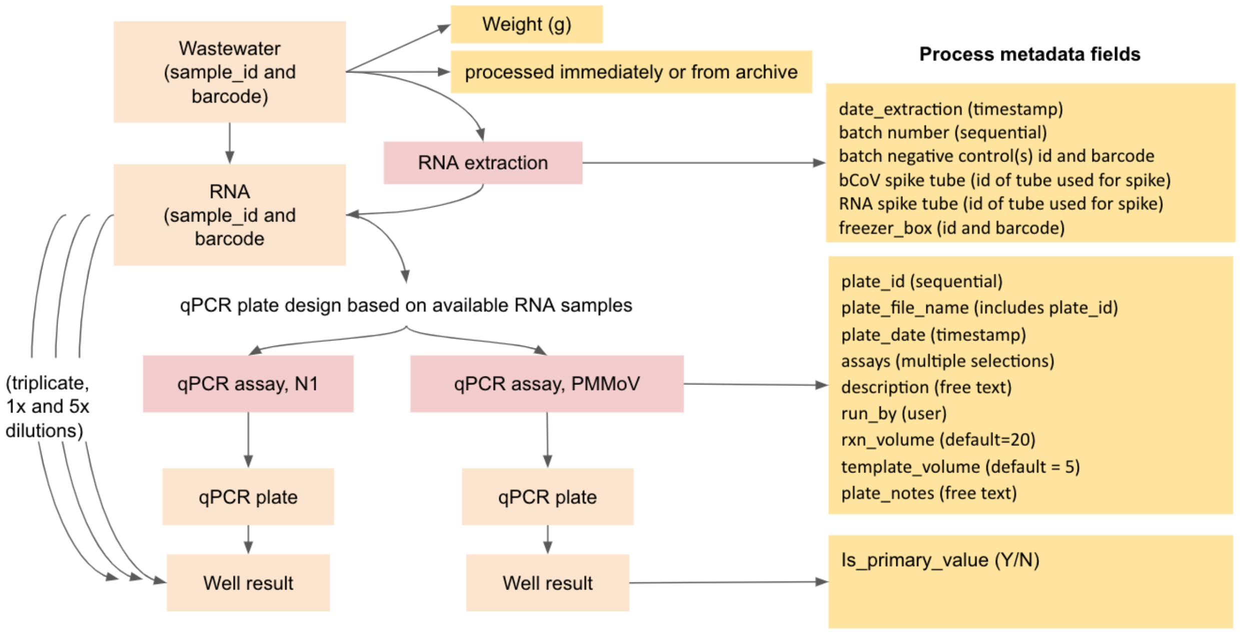

### Quality assurance and control plan

**UC Berkeley wastewater monitoring laboratory**

- **Sampling**
  - **Autosample failures:**
    - We log the total number of hours out of 24 hours represented in the 24-hr composite samples
    - We log any additional notes about samples including unusual events or observations made by field samplers
  - **Sample hold time:**
    - We overnight ship samples with an ice pack to the lab after salt has been added to preserve the RNA. We report sample hold time (time between retrieval of the composite sample and RNA extraction) and aim to keep this time ≤ 3 days.
- **RNA extraction**
  - **Negative controls:**
    - 40 mL of 1x PBS w/ 9.35g of NaCl, Tris HCl & EDTA, passed through extraction protocol (has bovine coronavirus and positive control RNA added. The negative controls should not contain SARS-CoV-2. Trace amounts of PMMoV in the negative control are permissible because PMMoV is highly concentrated in samples, and minor cross-contamination may occur.
  - **Positive controls:**
    - Lysis and recovery control: bovine coronavirus spiked into wastewater samples
      - Track which bovine coronavirus aliquot was spiked into each sample and save the original aliquots for later quantification alongside samples
    - RNA control: RNA spiked into wastewater samples
      - Track which RNA aliquot was spiked into each sample and save the original aliquots for later quantification alongside samples
      - All wastewater samples are spiked, but this assay is performed less frequently than bovine coronavirus
    - As an indicator of human fecal-associated viruses, we measure pepper mild mottle virus (PMMoV) in every sample. This virus should always be present at high concentrations, and low values may indicate extraction failure
  - **Replication:**
    - Biological replicate aliquots (2) are extracted from each 24-hour composite sample
  - **Contamination prevention:**

        - Before and after each extraction batch, biosafety cabinets, vacuum manifold, pipettes, and tube racks are cleaned with 10% bleach and 70% ethanol and UV irradiated for 15 minutes
        - Used labware and biohazardous waste are removed from the biosafety cabinet between extraction batches
    - **Reruns:**
        - Whole batches are excluded from reporting if the extraction negative control is positive for SARS-CoV-2 and is within 1 Cq of the highest sample in the batch (extremely rare); RT-qPCR may also be rerun to check the point at which the contamination occurred
        - Due to the extremely high concentration of PMMoV in wastewater samples, the extraction negative controls occasionally amplify for PMMoV at Cq ≥ 35. We do not consider this level of cross-contamination to be cause for concern as long as the concentrations in controls are several orders of magnitude lower than in samples.
        - Individual samples are excluded from reporting if the bovine coronavirus recovery is less than 1%. In this case, only the second biological replicate will be reported. If both replicates have poor recovery efficiencies, a third replicate will be extracted.

- **RT-qPCR**
    - **Negative controls:**
        - No-template controls included on every plate (3 wells per plate)
    - **Standards:**
        - Serially diluted standards included on every plate (3 replicates per point)
        - Standards are aliquoted upon delivery and stored at -80 C
        - Standards are DNA, which does not account for reverse-transcription efficiency but remains stable during storage
    - **Replication**
        - 3 technical replicate wells are analyzed for each RNA sample in each assay
    - **Template**
        - Template RNA is processed on the same day as extraction and never undergoes freeze-thaw (stored at 4 C between extraction and RT-qPCR)
        - Two aliquots of template RNA are stored at -80 C in case of sample reruns, and template is not freeze-thawed more than once
    - **Inhibition:**
        - for SARS-CoV-2 assays, RNA is assayed as 1x (undiluted) and 5x (1:4 dilution) of each sample and the highest effective concentration is reported (for PMMoV, only 10x diluted RNA is assayed)
    - **Plate preparation:**

        - To avoid data entry errors, pre-made templates are used within qPCR machine software and sample names are populated from sample inventory
    - **Contamination prevention:**
        - PCR work areas and pipettes are cleaned with 10% bleach, 70% ethanol and RNaseAway and UV irradiated for 15 minutes before and after each plate is prepared
        - Standards are aliquoted in a separate area to prevent contamination and false positives in samples
    - **Reruns:**
        - Whole plates are rerun if:
            - The efficiency of the standard curve does not fall between 90-110%
            - The no-template controls amplify
            - $R^2$ of standard curve is below 0.95

- **Data analysis**
    - The full data analysis pipeline can be found here: https://github.com/wastewaterlab/data_analysis
    - Outliers are removed from qPCR technical triplicates using Grubb's test (alpha = 0.05)
    - Raw data (RT-qPCR standard curves, controls, and samples) are visually inspected daily to identify outliers or potential errors in sample preparation or data entry
    - We have found that when all sample processing is consistent (same mastermix, primer/probe mixture, plates, and qPCR machine), using a single standard curve across all plates within one assay yields more consistent data than using the standard curve produced on each plate. Plate standard curves are used as quality control for qPCR efficiency before a default standard curve is applied to calculate concentrations in all samples on the plate.
    - All data are visualized in plots that are inspected in aggregate to identify any systematic patterns such as noticeably lower values for one plate, one day, or one week.
